# Increased Risk of Intracranial Hemorrhage in Older Patients Exposed to Multidrug Interactions Involving Warfarin

**DOI:** 10.1101/2023.09.27.23295040

**Authors:** Prathiv Raj Ramesh Babu

## Abstract

Multidrug interactions are a major cause of mortality for older patients. With an increase in “alert fatigue” for clinicians using Electronic Health Record systems (EHRs), Adverse Drug Events (ADEs) are increasing within older populations taking various drugs, because the probabilities of the adverse events associated with exposure to interacting drugs are not provided. The Observational Health Data Sciences and Informatics (OHDSI) ATLAS tool was utilized with Columbia University Irving Medical Center (CUIMC) patient data to determine whether Intracranial Hemorrhage (ICH) risk increases after being exposed to various multidrug interactions involving Warfarin in patients aged 60-90 years old one week before an ICH occurrence in patients prescribed ranges of 1-5, 6-10, and 11-15 drugs to also see the impact of polypharmacy on the prevalence of ICH amongst these patients. The prevalence of ICH in patients exposed to two and three-drug combinations involving Warfarin, Aspirin, Acetaminophen, and Amiodarone was measured. Significant increases in ICH risk and prevalence were identified in patients who had taken various combinations of these drugs with three-drug combinations involving Warfarin having a higher ICH risk than two-drug combinations involving Warfarin. Other findings also include that patients prescribed a larger number of drugs also tended to have a higher ICH risk than those prescribed a smaller number of drugs. Understanding the prevalence of such ADEs have crucial implications in the healthcare system, as the increased probabilities for certain ADEs from multidrug combinations can be applied to future EHR systems to counteract “alert fatigue” issues.

## Introduction

For older patients, drug-drug interactions are a major cause of mortality and morbidity [1]. When patients become older, the number of conditions they develop and the drugs they take increase, leading to an increase in potential multidrug interactions due to polypharmacy [1,2].

Polypharmacy increases the likelihood of two-drug interactions, but can also lead to multidrug interactions where a drug can interact with two or more other drugs. These interactions can lead to serious adverse drug events (ADEs) such as hemorrhage, serotonin syndrome, torsades de point, and seizures [2, 3]. In patients prescribed warfarin, known to interact with many drugs [4], these interactions can directly lead to gastrointestinal and intracranial hemorrhage (ICH).

There has been a widespread attempt to mitigate the negative effects of ADEs through drug interaction prompts in the Electronic Health Record (EHR). However, these prompts have been undermined by a rising trend in “alert fatigue” [3]. A recent study has estimated that approximately 5.5 million medication-related alerts were incorrectly avoided in the US alone at costs ranging from $871 million to 1.8 billion along with increased mortality and morbidity [5]. A reason that the alerts are avoided by clinicians is that the probabilities of ADEs are not reported when drug interaction alerts are provided. By finding the prevalence of ADEs within certain drug interactions, this issue could be counteracted by providing clinicians with the increase in the probability of an ADE for certain drug interactions as a function of the number of drugs that a patient is prescribed, taking polypharmacy into account in these calculations.This study focused on ICH, a class of hemorrhage classified as an ADE caused by drug interactions [3]. Intracranial Hemorrhage risk, especially in geriatric patients, has been found to increase significantly with the use of Warfarin [6]. ICH risk also increases with other Warfarin-interacting drugs in two-drug interactions like Aspirin [7], Acetaminophen [8], and Amiodarone [9]. The risk of ICH could be amplified in patients exposed to multidrug interactions involving three or more drugs, such as warfarin-aspirin and amiodarone, especially in patients exposed to polypharmacy. The objective of this retrospective cohort study was to measure the prevalence (PREV) of intracranial hemorrhage in patients exposed to multi-drug interactions involving warfarin (WA), aspirin (AS), acetaminophen (AC), and amiodarone (AM) and polypharmacy.

## Methods

### OHDSI and CUIMC Database

Within this study, patient data was sourced from New York Presbyterian Hospital (NYP), a major medical center in New York City that provides inpatient and outpatient care. The Observational Health Data Sciences and Informatics (OHDSI) CUIMC database contains data from the EHRs of 4,618,530 patients from 1985 onwards. The database contains information regarding patient demographics, conditions, drugs prescribed, measurements (vital signs and laboratory tests), devices, and other observations from healthcare providers. Data sources include current and previous EHR systems along with other administrative systems as well.

### Mining Data on OHDSI

Anonymous patient data stored in OHDSI’s Observational Medical Outcomes Partnership (OMOP) model is accessible to researchers through ATLAS, an open-source tool that searches for relevant data utilizing structured queries. On ATLAS, a researcher can ask (for example) “How many patients aged 60-90 with a Myocardial Infarction have been prescribed Statins?” Through ATLAS queries the association between Intracranial Hemorrhage and various multidrug combinations involving Warfarin was explored.

### Patient Counts on certain Multidrug Combinations with and without Intracranial Hemorrhage

ATLAS, OHDSI’s SQL tool, can count the number of patients within a cohort that meet specific inclusion and exclusion criteria. Two cohorts were created to conduct this study, the ICH COHORT (Figure 1), which was designed to capture the number of patients with an Intracranial Hemorrhage on Warfarin (WA), and combinations of Warfarin-interacting drugs including Aspirin (AS), Acetaminophen (AC), and Amiodarone (AM).

**Figure 1.**
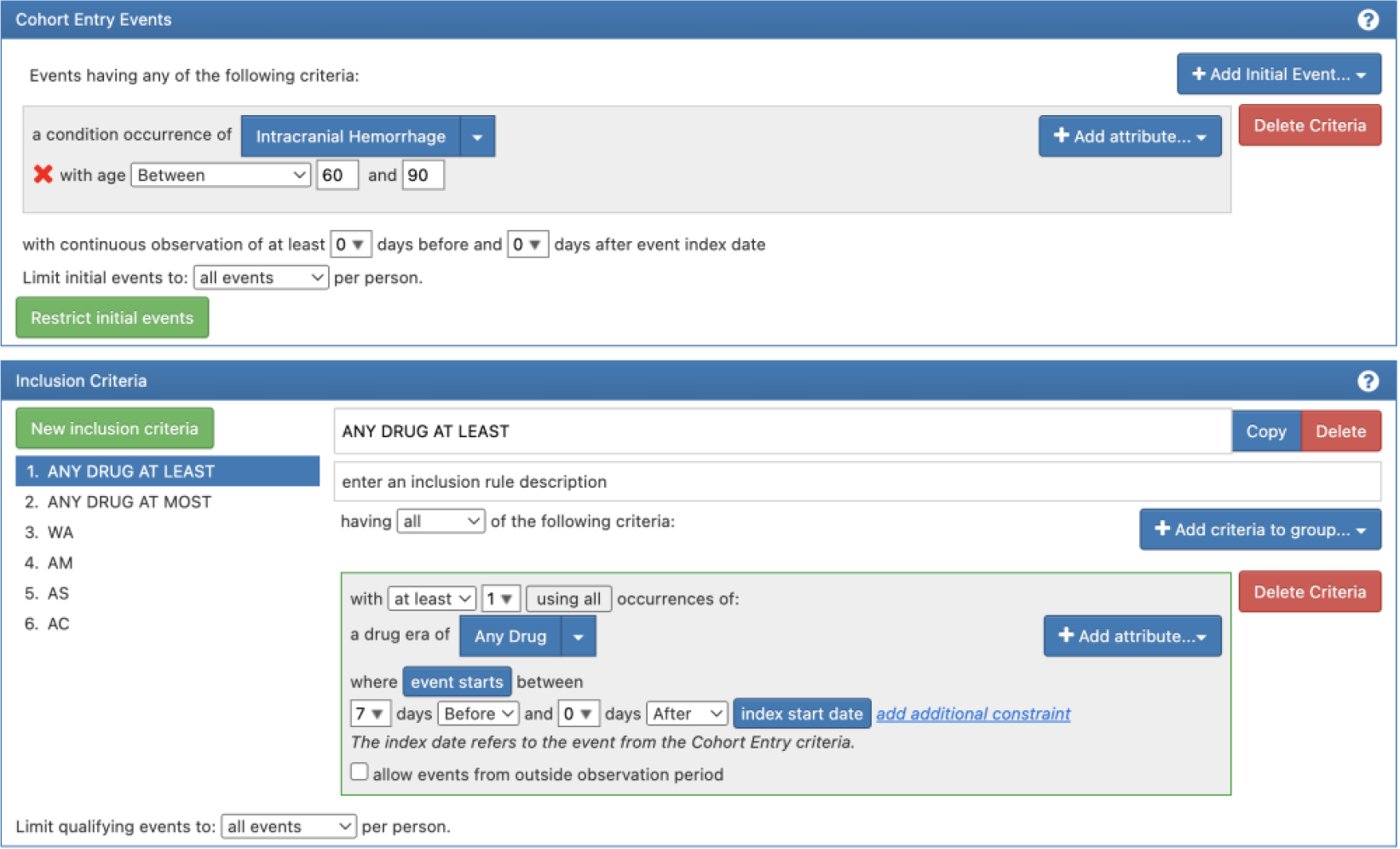
ICH Cohort Entry and Inclusion Settings including ICH Occurrence, Drugs of Interest (Warfarin, Amiodarone, Aspirin, & Acetaminophen), and ranges of “any drugs” prescribed to patients.

Patients with the Intracranial Hemorrhage condition on OHDSI were identified through SNOMED-CT code 1386000 and all the included descendants of that code (subtypes of Intracranial Hemorrhage). The Cohort Inclusion Criteria provided the total number of patients with or without a prescription for one or more of the drugs of interest seven days before an Intracranial Hemorrhage. The Inclusion Criteria also contained the total number of drugs a patient was prescribed in ranges of [1-5], [6-10], or [11-15].

The second DRUG COUNTS COHORT (Figure 2) was designed to capture the number of patients with a prescription for two or more drugs on the same day, regardless of whether they had an Intracranial Hemorrhage.

**Figure 2.**
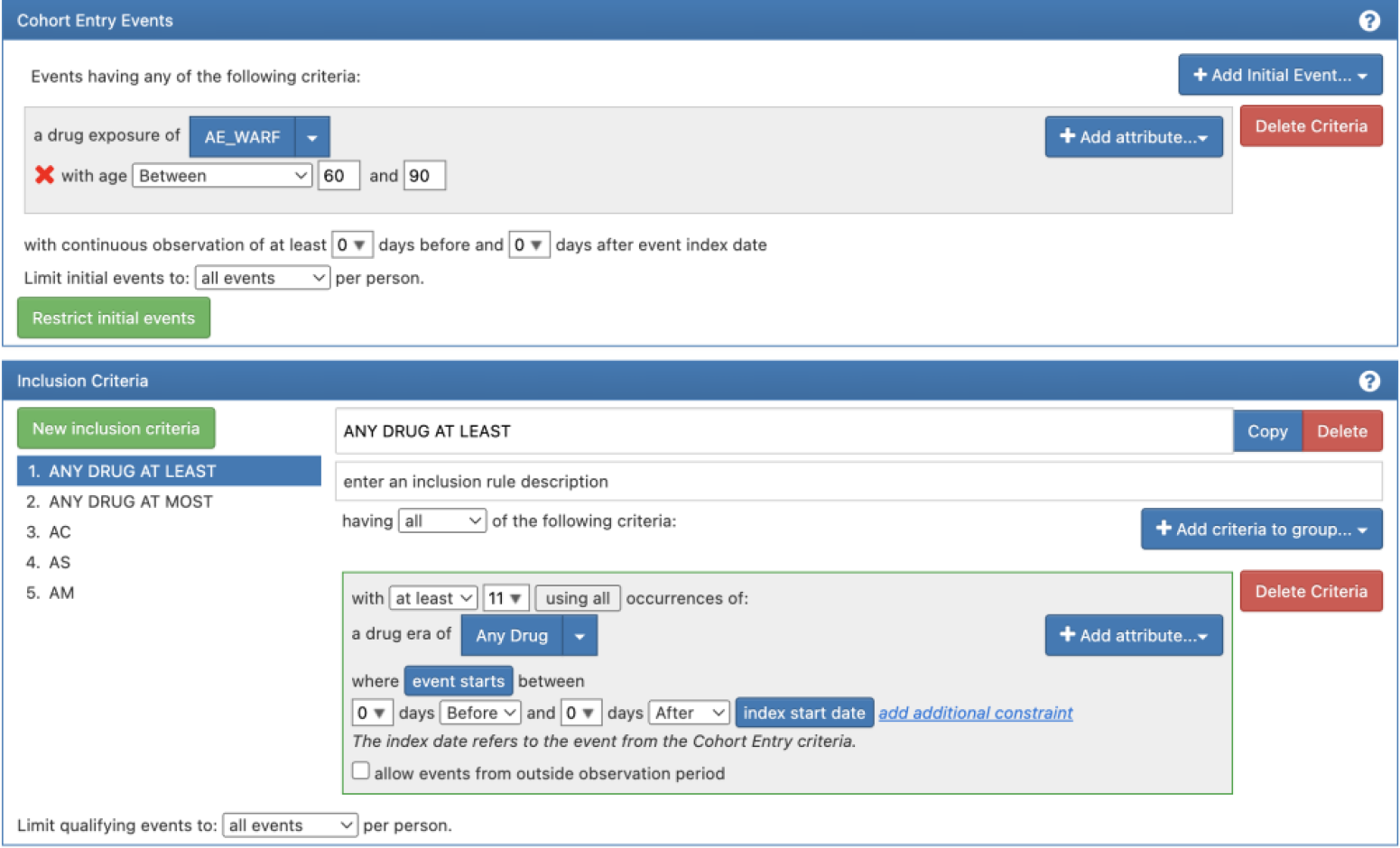
Drug Counts Cohort Entry and Inclusion Criteria including exposures to Drugs of Interest (Warfarin, Amiodarone, Aspirin, & Acetaminophen), and ranges of “any drugs” prescribed to patients.

Cohort Inclusion Criteria provided the total number of patients with or without a prescription for two or more of the drugs (including Warfarin) of interest on the same day, regardless of ICH occurrence. The Inclusion Criteria also contained the total number of drugs a patient was prescribed in ranges of [1-5], [6-10], or [11-15]. Drugs were found through their respective RxNorm IDs. All variations and doses of these drugs were included in search queries.

### Calculation of ICH Prevalence and Risk

ICH prevalence (Prevalence [ICH]) was calculated using the number of patients with an Intracranial Hemorrhage on certain drug combinations and the number of patients across the entire CUIMC OHDSI database who were prescribed those respective drugs.

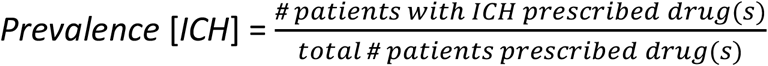

Utilizing ICH Prevalence, the Relative Risk (RR) of ICH was calculated in patients prescribed both three drugs in a multidrug combination compared to patients prescribed two out of the three drugs in the aforementioned combination.

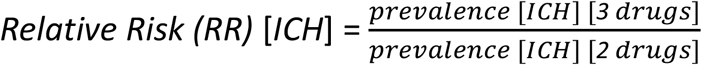

Standard 95% Confidence Intervals for RR were also calculated using conventional statistical methodology.

### Drugs focused on within the Study

As mentioned earlier, a focus was established on Warfarin-interacting drugs that have shown increases in the risk of Intracranial Hemorrhage, like Aspirin [7], Acetaminophen [8], and Amiodarone [9].

### Polypharmacy Influence on ICH Risk

To study the relationship between the prevalence of ICH and the number of drugs the patients were prescribed, the cohorts were divided into three groups based on the number of drugs prescribed: [1-5], [6-10], or [11-15] drugs.

### Other Statistics

The prevalence of Intracranial Hemorrhage between patients prescribed both drugs in a certain multidrug combination was compared to the prevalence of Intracranial Hemorrhage amongst patients prescribed without the second in that two-drug combination utilizing the conventional test for significance between two independent proportions. Proportions were considered significantly different if p < 0.01, with p values originating from two-tailed analyses. Microsoft Excel was utilized for these calculations.

## Results

### Patient Population

The OHDSI CUIMC Database consists of 1,015,532 patients aged 60 to 90 whose EHR noted at least one condition. Of this group, 529,271 patients had at least one drug prescribed during any one of their interactions with CUIMC and its associated health system. 14,281 patients (1.41%) had a SNOMED-CT code recorded for an Intracranial Hemorrhage at some point during an inpatient or outpatient visit.

### Drug Exposure

Of the 529,271 patients 33,527 (6.3%) were prescribed Warfarin. A significant proportion of patients were prescribed Aspirin (33.6%) or Acetaminophen (44.4%), while a much lower proportion were prescribed Amiodarone (3.8%). Of the patients prescribed Warfarin, 39.1% were prescribed Aspirin, and 51.1% were prescribed Acetaminophen. A substantial number of patients were exposed to three interacting drugs: 22% of patients prescribed WA were also prescribed AS and AC, and > 50% of patients prescribed WA and AM were also prescribed AS or AC.

### Statistical Findings

Significant findings included that the Prevalence of ICH is significantly higher in patients exposed to three interacting drugs rather than just two (Figure 3).

**Figure 3.**
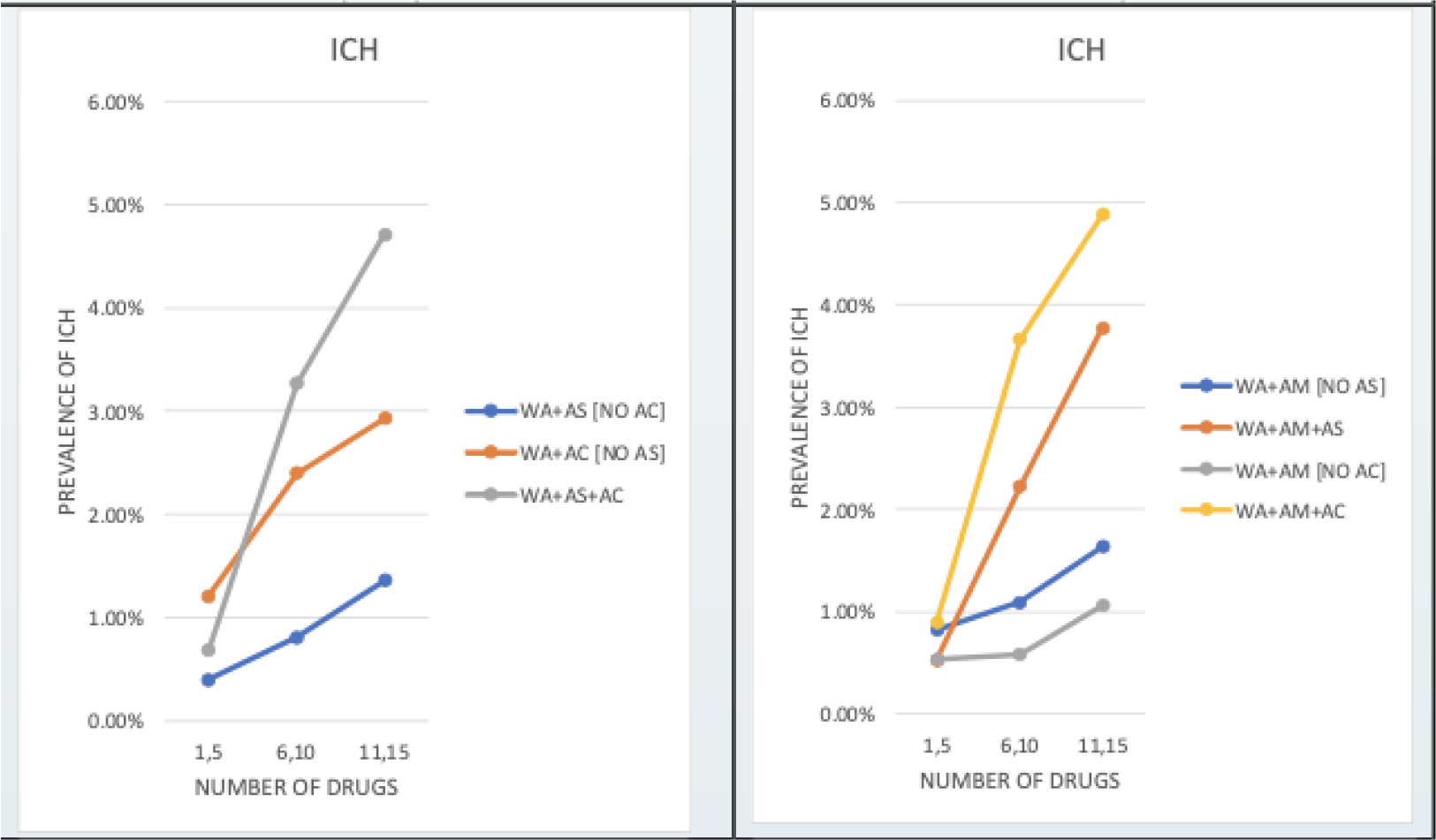
Intracranial Hemorrhage Prevalence across Multidrug Interactions involving Warfarin.

As the total number of drugs prescribed increases from [1-5] to [11-15], the prevalence of ICH across each combination subsequently increases as well. The results also depict an increase in prevalence when observing multidrug combinations (e.g. WA, AM, and AC) in comparison to two-drug combinations without the third drug in the multidrug combination (e.g. WA, AM, no AC). Significant increases in prevalence include an approximately 3.5 times increase between WA and AS while prescribed 11-15 drugs and WA, AS, and AC while prescribed 11-15 drugs.

Combinations involving AC tended to have a higher ICH prevalence than those with the same number of drugs but without AC. ICH prevalence for WA and AC, for example, was higher than the ICH prevalence for WA and AS across all three drug ranges. Even with Amiodarone in the multidrug interactions, the ICH prevalence for WA, AM, and AS was lower than the ICH prevalence for the WA, AM, and AC interaction.

These trends also roughly applied to Relative Risk findings (Table 1).

**Table 1.** Relative Risk of Intracranial Hemorrhage of patients on three drugs (including Warfarin) over patients on two drugs (including Warfarin).

| ICH | RELATIVE RISK |  |  |
| --- | --- | --- | --- |
| WA+ | # "any drug" |  |  |
|  | 1-5 | 6-10 | 11-15 |
| AS+AC versus -AS-AC | 1.06 | 4.42 | 7.72 |
| AM+AS versus +AM-AS | 0.64 | 2.03 | 2.31 |
| AM+AC versus +AM-AC | 1.67 | 6.28 | 4.59 |

Relative Risk tended to generally increase as patients were prescribed more drugs, especially with drastic increases across all three drug prescription ranges for WA, AS, and AC compared to WA without AS or AC.

However, some interesting findings were discovered within the Amiodarone multidrug combinations. WA with AM and AC versus WA with AM and without AC found its highest Relative Risk to be within the 6-10 total drugs prescribed range with a decrease when the range of prescribed drugs increased to 11-15. The multidrug interaction with WA, AM, and AS even yielded a lower Relative Risk within the 1-5 drugs prescribed range than the two-drug interaction of just WA and AM without AS.

Regarding other statistical results, there were also significant differences in the prevalences (proportions) of ICHs when comparing interactions with three drugs compared to two drugs.

## Discussion

### Electronic Health Record Applications

According to the aforementioned findings, the PREV of ICH was higher in patients exposed to three interacting drugs compared to two. The PREV and RR of ICH were lowest in patients prescribed 1-5 drugs and highest in those prescribed 11-15 drugs.

While these results show the significant repercussions of these drug-drug interactions, most electronic health record (EHR) systems alert providers regarding these interactions, but these alerts are generally avoided due to “alert fatigue” [3]. Improving the alert system to have clinicians accurately report these interactions could have a widespread impact on the healthcare system, considering how frequently these drug alerts are avoided.

Overriding of the drug-drug interaction warning prompts might be reduced if the prompt was fired only for the high prevalence multidrug interactions and RR > 5 prescribed drugs. Even solutions like these can greatly reduce the ignorance of these alerts, which carry significant repercussions with a cost projection of $871 million to $1.8 billion [5] for treating preventable adverse drug events (like intracranial hemorrhages) in the United States.

### Limitations

A primary limitation of this study was that it was based on data from a singular medical center.

While it was assumed that ICH risk and prevalence increased as patients took more drugs, it is imperative to note that the OHDSI CUIMC database solely notes that the drug has been prescribed to the patient, not taken. Current conclusions can only state that the increased risk was due to the prescription of the drug rather than the effects of the usage of the drug itself.

Along with this, in larger ranges of drugs prescribed (beyond 5), the increase in risk could also be a result of drug-disease interactions. This means that the increase in risk could be due to the way a certain drug interacts with another disease or condition a patient may have, contributing to the development of an Intracranial Hemorrhage.

A factor involved in these findings could be that patients on one combination of drugs may not be the same as the patients on another combination of drugs. Other factors like other conditions they may possess could also play a role in increasing the prevalence or the risk of developing an Intracranial Hemorrhage across comparison groups.

### Future Work

Regarding whether patients on one combination of drugs are essentially the same as the patients on another combination of drugs, a Propensity Score Matching study can be conducted to reduce the effects of confounding within a study [10]. The procedures within this study can also be expanded to larger global healthcare databases to get a larger sample size of patients.

To directly establish causation between a patient taking a certain combination of drugs and an increase in Intracranial Hemorrhage prevalence, a clinical study could be administered with a similar setup as that analyzed by Geoffrey C. Cloud [11] in which patients on certain drug combinations are observed and tracked for Intracranial Hemorrhage occurrences after drug administration.

## Conclusion

Geriatric patients are exposed to a variety of drug-drug interactions, including those that can cause morbidity and mortality in these patients [1]. Within this study, OHDSI’s CUIMC database was utilized to understand whether Intracranial Hemorrhage prevalence and risk increases following various multidrug interactions involving Warfarin. The discovery of Intracranial Hemorrhage Prevalences and Risk Increases are extremely pivotal for Electronic Health Records. Firing a drug interaction prompt only for the high prevalence multidrug interactions and RR > 5 prescribed drugs can take significant steps towards reducing clinician alert avoidance and reducing the large number of clinician alert overrides along with the the large medical costs associated with them.

## Acknowledgments

Thanks to Dr. Herbert S. Chase Jr. for his mentorship throughout this study within Columbia University’s Department of Biomedical Informatics Summer Fellowship Program along with Dr. Soojin Park for providing a spark of interest in Intracranial Hemorrhage research. Thanks as well to Dr. Matthew Spotnitz and Dr. Eugene Kim for assisting with OHDSI inquiries throughout the study, along with Mr. Kang-Chih Chen and Dr. Karthik Natarajan for assisting with technological setup on Columbia’s OHDSI platform.

## Funding Acknowledgement

The author received no financial support for the research, authorship, and/or publication of this article.

## Ethics Declaration

Research involving human data was performed in accordance with the Declaration of Helsinki and was approved by the Columbia University Irving Medical Center Institutional Review Board under the protocol AAAU1902 “Multidrug Interactions”. Obtaining consent was waived by the CUIMC Institutional Review Board given that it would not be feasible to obtain consent on the thousands of patients used in the study and that using the data posed no risk to the patients.

## Data Availability

Due to HIPPA regulations, supporting data cannot be made openly available. For further information about the data or access conditions please contact Herbert S. Chase Jr. via

## References

1. Błeszynska E, Wierucki Ł, Zdrojewski T, Renke M. Pharmacological interactions in the elderly. Medicina (Kaunas). 2020 Jun 28;56(7):320.

2. Björkman IK. Drug-drug interactions in the elderly. The Annals of Pharmacotherapy. 2002;36(11):1675–81.

3. Anand TV, Wallace BK, Chase HS. Prevalence of potentially harmful multidrug interactions on medication lists of elderly ambulatory patients. BMC Geriatr. 2021 Nov 19;21(1):648.

4. Holbrook AM, Pereira JA, Labiris R, et al. Systematic overview of warfarin and its drug and food interactions. Arch Intern Med. 2005 May 23;165(10):1095–1106.

5. Slight SP, Seger DL, Franz C, Wong A, Bates DW. The national cost of adverse drug events resulting from inappropriate medication-related alert overrides in the united states. J Am Med Inform Assoc. 2018 Sep 1;25(9):1183–1188.

6. Jimenez-Ruiz A, Gutierrez-Castillo A, Ruiz-Sandoval JL. Fatal Intracranial Hemorrhage Associated with Oral Warfarin Use. Cureus. 2018 Nov 12;10(11):e3571.

7. Hart RG, Benavente O, Pearce LA. Increased risk of intracranial hemorrhage when aspirin is combined with warfarin: A meta-analysis and hypothesis. Cerebrovasc Dis. 1999 Jul-Aug;9(4):215–7.

8. Lopes RD, Horowitz JD, Garcia DA, Crowther MA, Hylek EM. Warfarin and acetaminophen interaction: a summary of the evidence and biologic plausibility. Blood. 2011 Dec 8;118(24):6269–73.

9. Lam J, Gomes T, Juurlink DN, Mamdani MM, Pullenayegum EM, Kearon C, Spencer FA, Paterson M, Zheng H, Holbrook AM. Hospitalization for hemorrhage among warfarin recipients prescribed amiodarone. Am J Cardiol. 2013 Aug 1;112(3):420–3.

10. Austin PC. An introduction to propensity score methods for reducing the effects of confounding in observational studies. Multivariate Behav Res. 2011 May;46(3):399–424.

11. Cloud GC, Williamson JD, Thao LTP, et al. Low-dose aspirin and the risk of stroke and intracerebral bleeding in healthy older people: secondary analysis of a randomized clinical trial. JAMA Netw Open. 2023;6(7):e2325803.

